# Individuals with knee osteoarthritis show few limitations in reactive stepping responses after gait perturbations

**DOI:** 10.1101/2023.10.04.23296525

**Authors:** R.J. Boekesteijn, N.L.W. Keijsers, K. Defoort, A.C.H. Geurts, K. Smulders

## Abstract

**Background:** Knee osteoarthritis (OA) causes structural joint damage. The resultant symptoms can impair the ability to recover from unexpected gait perturbations, contributing to an increased fall risk. This study compared reactive stepping responses to gait perturbations between individuals with knee OA and healthy individuals.

**Methods:** Kinematic data of 35 individuals with end-stage knee OA, and 32 healthy individuals in the same age range were obtained during perturbed walking on a treadmill at 1.0 m/s. Participants received anteroposterior (trip or slip) or mediolateral perturbations during the stance phase. Changes from baseline in margin of stability (MoS), step length, step time, and step width during the first two steps after perturbation were compared between groups using a linear regression model. Extrapolated center of mass (XCoM) excursion was descriptively analyzed.

**Findings:** After all perturbation modes, XCoM trajectories overlapped between individuals with knee OA and healthy individuals. Participants predominantly responded to mediolateral perturbations by adjusting their step width, and to anteroposterior perturbations by adjusting step length and step time. None of the perturbation modes yielded between-group differences in changes in MoS and step width during the first two steps after perturbation. Small between-group differences were observed for step length (i.e. 2 cm) of the second step after trip and slip perturbation, and for step time (i.e. 0.02 s) of the second step after slip perturbations.

**Interpretation:** Despite considerable pain and damage to the knee joint, individuals with knee OA showed comparable reactive stepping responses after gait perturbations to healthy participants.

## 1. Introduction

Knee osteoarthritis (OA) is a debilitating joint disease characterized by degradation of articular cartilage and structural damage to the knee joint (1). Common symptoms of knee OA include pain, stiffness, knee instability, muscle weakness, and fatigue. In addition, knee OA may lead to afferent and efferent neural deficits, expressed by reduced vibratory sense (2), reduced proprioception (3), and poorer control over muscle force generation (4). These symptoms could lead to impaired stability during walking in individuals with knee OA (5). Indeed, observational studies suggest that individuals with knee OA are 25-54% more likely to experience a fall compared to those without knee OA (6–10).

Stable gait can be defined as “gait that does not lead to falls in spite of perturbations” (11). The application of unexpected, external perturbations to challenge gait stability has become a common method to study dynamic balance control in humans (12–17). To ensure adequate recovery from such external perturbations, the body’s extrapolated center of mass (XCoM) – which is the center of mass (CoM) position plus its velocity vector divided by the inverted pendulum’s eigenfrequency (18) – needs be controlled with respect to the limits of a continuously changing base of support (BoS). This process relies on the integration of diverse sensory inputs into an adequate motor response. Dynamic balance control is believed to be actively regulated, particularly in the mediolateral (ML) direction (19), whereas in the anteroposterior (AP) direction, it may be relatively less controlled (20) due to exploitation of passive system dynamics (21). Three main mechanisms can be used to actively regulate AP and ML gait stability during walking: 1) foot placement, 2) changing the position of the center of pressure under the stance foot, and 3) modulating the body’s angular momentum (22). Among the three mechanisms, foot placement is considered the most dominant (23).

To study the effects of knee OA on gait stability, responses to AP (24–27) and ML (25, 28, 29) gait perturbations have previously been compared between individuals with knee OA and healthy participants. Overall these studies showed mixed results, with some showing effects of knee OA on perturbation responses (24–27) and others finding no such effects (28, 29). Outcomes of these studies included muscle activation (including quadriceps, hamstrings, calf muscles) (25, 28, 29), lower-extremity kinematics (24–29), lower-extremity kinetics (24), and step characteristics (24, 26, 27). However, none of the studies investigated gait stability as the relationship between CoM state and foot placement. Furthermore, because stability measures vary with differences in gait speed (30, 31), and gait speed of individuals with knee OA is lower compared to healthy individuals (32), gait speed should be considered as a confounder in these comparisons. Unfortunately, none of the previous studies (24–29) controlled for gait speed in their experiments.

In this study, we examined reactive balance responses to ML and AP perturbations in individuals with knee OA, and compared them to responses of healthy peers walking at a predefined, fixed speed. We hypothesized that, compared to healthy participants, individuals with knee OA would show larger XCoM excursions after perturbation, leading to a lower MoS in the first step after both ML and AP perturbations.

## 2. Methods

### 2.1 Participants

This study was part of a longitudinal study investigating real-life and challenging gait skills in individuals scheduled for total knee arthroplasty (TKA) (https://osf.io/64ejm). Real-world gait data of this study has been published as preprint (33). Thirty-five individuals with end-stage knee OA, scheduled for cruciate retaining TKA, and thirty-two healthy controls (HC) participated in this study. Individuals with knee OA, who were candidates for posterior cruciate retaining TKA at the Sint Maartenskliniek Nijmegen, were screened by a research nurse for eligibility. Eligibility criteria included: 1) symptomatic and radiological knee OA (i.e. Kellgren-Lawrence grade > 2), 2) intact posterior cruciate ligament, 3) correctable or <10° rigid varus or valgus deformity of the knee, and 4) stable health (ASA-score ≤ 3), 5) aged between 40-80 years. Healthy participants were recruited from the community, in the same age range and with similar sex distribution as the group of individuals with knee OA. Healthy participants were matched to the individuals with knee OA that received the Journey II CR implant (Smith & Nephew, Memphis, TN, USA) based on age and sex (which was the case for 32 out of 35 participants), allowing a maximum age difference of 5 years. Healthy participants had no diagnosis of knee OA and had no self-reported pain complaints in the lower-extremities. Exclusion criteria for both groups were: 1) BMI > 35 kg/m^2^, 2) moderate to severe knee, hip or ankle pain defined as an average score >4 on items 3-6 of the Short Brief Pain Inventory; excluding the knee indexed for TKA, 3) previous knee, hip, or ankle joint replacement, 4) any other musculoskeletal, neurological, or uncorrected visual disorder impairing gait or balance. Informed consent was obtained from all participants prior to the experiments. Ethical approval was obtained from the CMO Arnhem/Nijmegen (2019–5824). All study methods were carried out in accordance with the Declaration of Helsinki.

### 2.2. Clinical assessments

AP X-rays, available through regular clinical care, were scored by KD using the Kellgren and Lawrence grades (34). Anthropometric characteristics (height, body mass, and BMI) were obtained on the same day as the gait assessment. For individuals with knee OA, this was on average 1.8 months (IQR = 1.5) before TKA. All participants reported pain scores during activity and rest using a numeric rating scale (NRS). In addition, the Knee injury and Osteoarthritis Outcome Score – Physical Function shortform (KOOS-PS) (35) and the clinical and functional score of the Knee Society Score (KSS) (36) were obtained for individuals with knee OA. Fall history was assessed by asking the participants if they had experienced a fall during the 3 months preceding the study visit (37). If participants reported they had fallen, the number of falls was recorded.

### 2.3 Equipment

Participants walked on an instrumented split-belt treadmill (GRAIL, Motek Medical BV, The Netherlands) that was surrounded by a 180° semi-cylindrical screen with a virtual environment. For safety reasons, all participants wore a safety harness when walking on the treadmill. Participants were equipped with twenty-three reflective markers, following the Vicon Lower Body model (38), with additional markers placed on C7, and bilaterally on the acromion process, humeral lateral epicondyle, and the ulnar styloid process. These additional markers were used to account for trunk and arm movements in the CoM estimation (39). Marker data were acquired using a ten-camera motion capture system (Vicon, Oxford, UK).

### 2.4 Procedures

Participants were first familiarized with the experimental set-up, including walking on the treadmill with virtual environment. Subsequently, comfortable walking speed was determined using the protocol described in Hak *et al.* (40), which started at a speed of 0.5 m/s with increments or decrements of 0.05 m/s. The perturbation protocol consisted of two separate sequences with ML and AP perturbations (Figure 1C). These sequences consisted of a block of perturbations of approximately 3 minutes, which was repeated twice with 2 minutes of rest in between. During these sequences, walking speed was fixed at 1.0 m/s, which was based on the mean overground comfortable walking speed of individuals with knee OA (e.g. 0.97 m/s; SD = 0.17 (41)) as well as by pilot testing.

**Figure 1:**
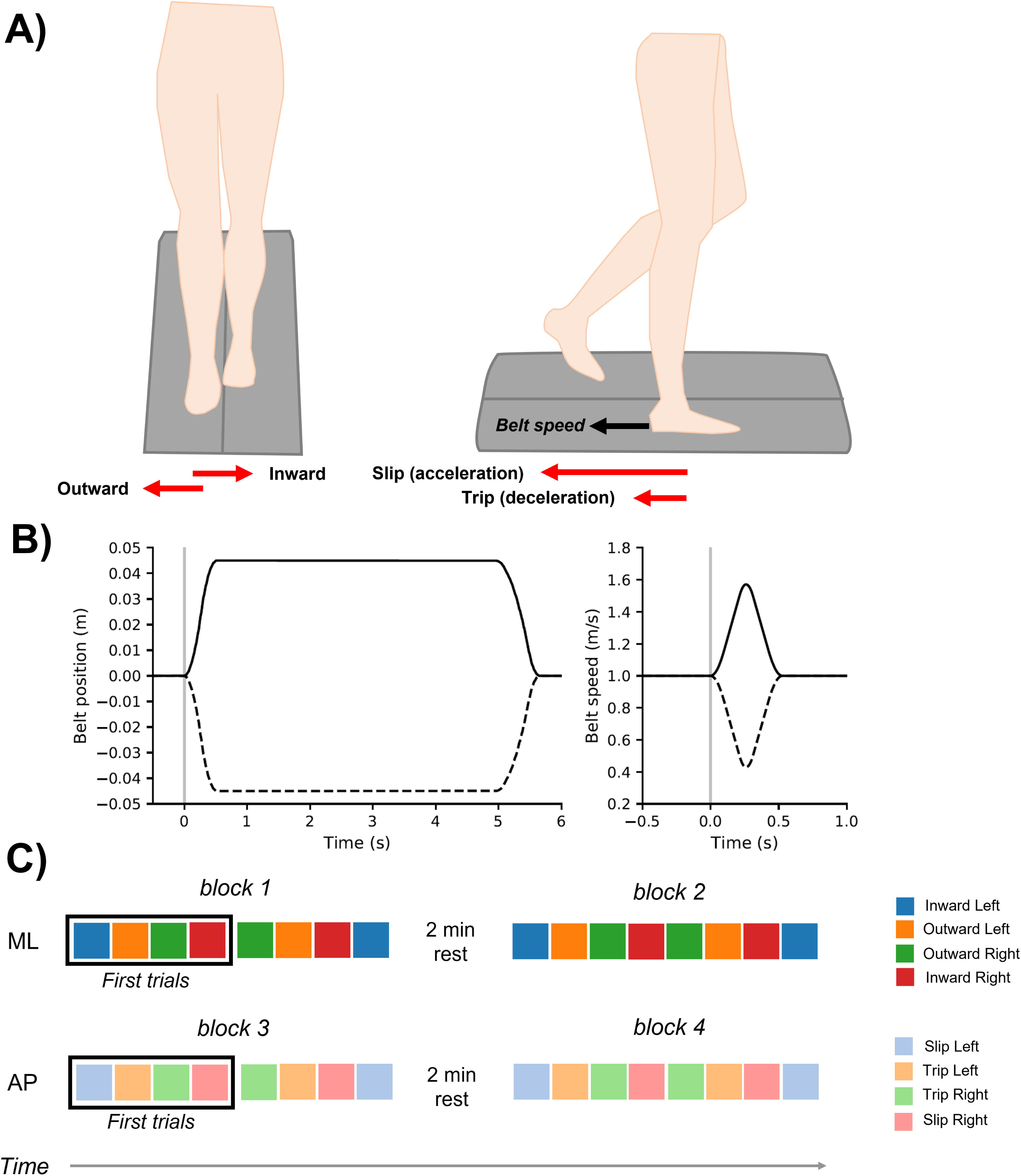
Overview of the experimental design. Definitions of the different perturbation modes are provided in panel A. Panel B shows the perturbation profiles for mediolateral and anteroposterior perturbations. T=0, indicated by the gray line, corresponds to the heel contact of the perturbed leg. In panel C, the design of the mediolateral and anteroposterior perturbation sequences are shown, with each color representing a unique perturbation mode.

Perturbations consisted of 4.5 cm platform translations in 0.5 s (ML) or a change in belt speed with a speed difference of 0.6 m/s in 0.5 s (AP). For ML perturbations the platform always returned to the middle, neutral position 5 seconds after initial perturbation, which was necessary as total platform movement was limited to 5 cm at each side. Perturbations were triggered by heel contact and delivered during the stance phase (Figure 1B).

There were four different perturbation modes for each sequence (ML vs. AP), depending on side (i.e. affected vs. unaffected in patients and left vs. right in healthy participants) and direction (i.e. inward vs. outward in ML perturbations, and slip vs. trip in AP perturbations; Figure 1A). The definition of side in healthy participants was matched to the affected side of an individual with TKA with similar sex and age. Each of 4 perturbation modes were repeated twice within a block, resulting in a total of 8 perturbations per block (Figure 1C). The order of perturbations was fixed, but concealed to the participants. To prevent carry-over effects, the duration between two consecutive perturbations was at least 7 seconds. The exact interval between perturbations varied in order to prevent anticipation.

### 2.5 Outcomes and data analysis

Data were processed in Octave 6.3.0 and figures were prepared in Python 3.8.3. Marker data were filtered using a 2^nd^ order low-pass Butterworth filter with a cut-off frequency of 10 Hz. Gait events were detected using the velocity-based algorithm described by Zeni Jr. *et al.* (42). From marker data, the CoM position was determined using the methods described by Tisserand *et al.* (39). Subsequently, the XCoM was calculated based on the inverted pendulum model, using the formula presented by Hof

*et al.* (18):

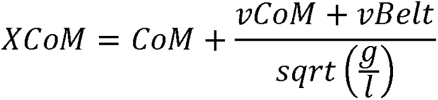

*where XCoM is the body’s extrapolated center of mass, CoM the CoM position, vCoM the CoM velocity, vBelt the belt speed (1.0 m/s for the anteroposterior direction), g the gravitational acceleration (9.81 m/s^2^), and l is defined as the pendulum height (height of the CoM)*.

To descriptively analyze CoM and XCoM, trajectories were time normalized from the second step before perturbation until the fifth step after perturbation. In addition, CoM position at heel strike before perturbation was subtracted from the entire time series, such that group averages could be taken. The MoS was calculated separately in the ML and the AP direction (Figure 2). For the AP direction, MoS was calculated as the difference between the toe marker and XCoM at heel strike. For the ML direction, MoS was calculated as the minimum of the difference between the ankle marker and XCoM position during stance, which was approximately at the instant of opposite toe-off (18). Positive MoS values indicate instantaneous stability, whereas negative MoS values indicate instantaneous instability. Discrete parameters (MoS, step time, step length, and step width) were calculated for the three steps before each perturbation (i.e. step-2, step-1, and pre) until five steps after perturbation (i.e. post1 – post5). Step length was defined as the difference in AP position of the heel markers between two consecutive heel strikes, plus step time times belt speed. Step width was defined as the difference in ML position of the heel markers between two consecutive heel strikes. For both step length and step width calculations, we accounted for changes in belt speed or platform translation, such that these parameters included the distance from the perturbation. First repetitions of each perturbation mode were removed from analysis, as they may elicit inherently different responses than later repetitions (e.g. due to first trial effect; (43)). In addition, all responses during which the handrail was touched were removed from analysis. Touching of the handrail was visually identified by the investigator.

**Figure 2:**
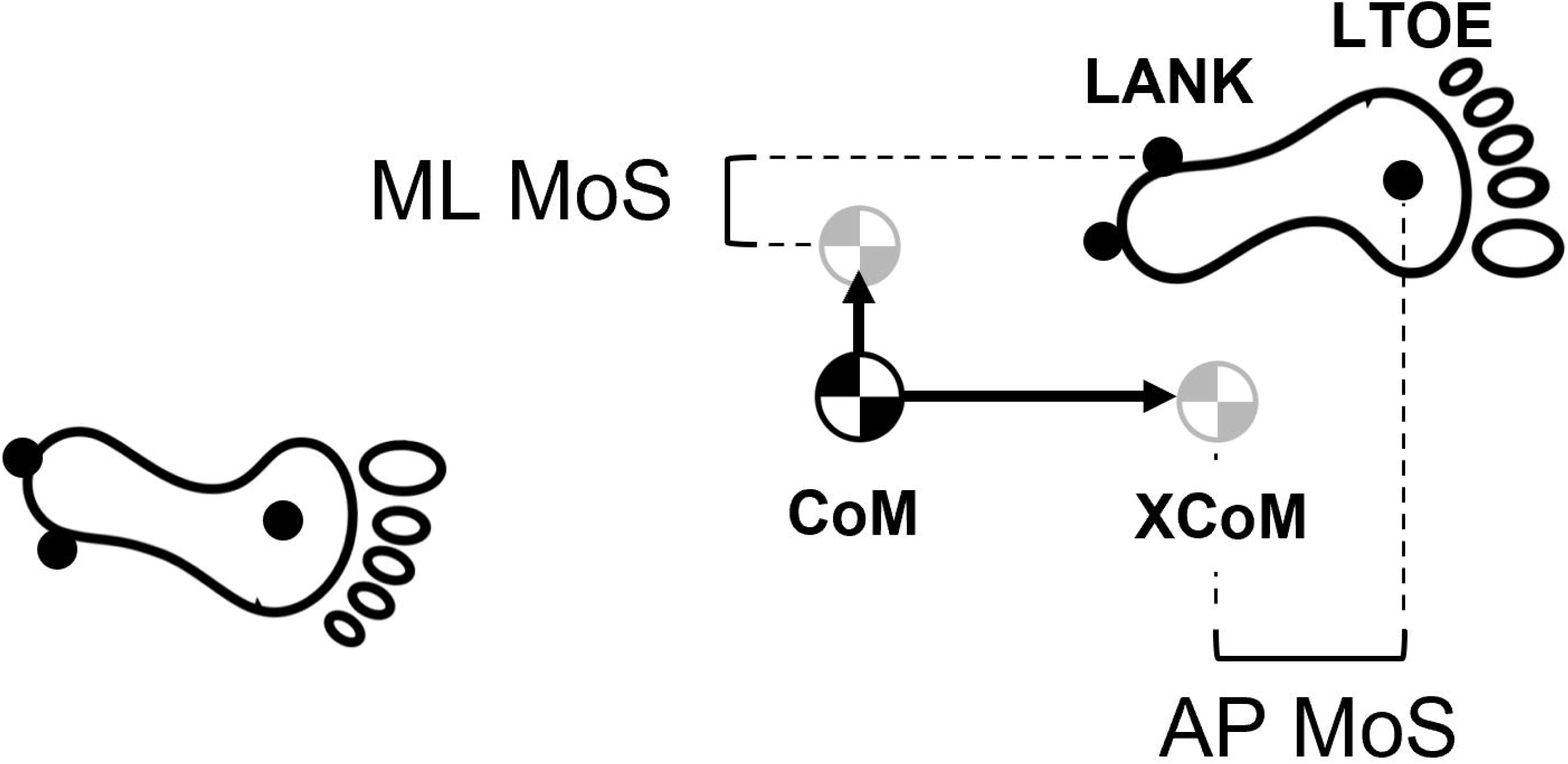
Simplified overview of the definition of the margin of stability in the ML and AP direction. AP MoS was calculated in the forward direction, meaning that AP MoS is positive when XCoM was behind the toe marker.

### 2.6 Statistical analysis

To reduce the risk of type I errors, between-group effects were only tested in the first two steps after perturbation (i.e. post-1 and post-2). For similar reasons, we only compared data of perturbations to the affected leg between groups, as the largest differences could be expected here. The two steps before each perturbation trial (step-2 and step-1) were combined into a baseline score to reduce noise and average out potential asymmetries. For each outcome measure, two separate linear regression models were created, with difference from baseline as the dependent variable (ΔY_post1/2_), group as independent variable, and baseline score (Y_baseline_) as covariate:

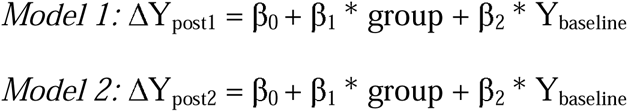

In these models Y was the variable of interest (i.e. MoS, step width, step length, or step time). Between-group differences (i.e. β_1_ derived from the models) were reported as mean differences with 95% confidence intervals. Furthermore, changes over time (i.e. ΔY_post1_ and ΔY_post2_) were estimated. If there was no significant group effect (p > 0.05), the factor group was removed from the statistical model to estimate ΔY_post1/2_ for all participants. XCoM and CoM trajectories were descriptively analyzed. Statistical analysis was performed in RStudio using the stats package (version 4.1.2).

## 3. Results

Baseline characteristics are provided in Table 1. Individuals with knee OA had a higher body mass, higher BMI, and experienced more pain during activity and rest compared to healthy controls. Comfortable walking speed was -0.21 m/s lower in individuals with knee OA than in healthy controls. Four participants with knee OA (11%) and two healthy participants (6%) reported they had fallen during the preceding 3 months.

**Table 1:**
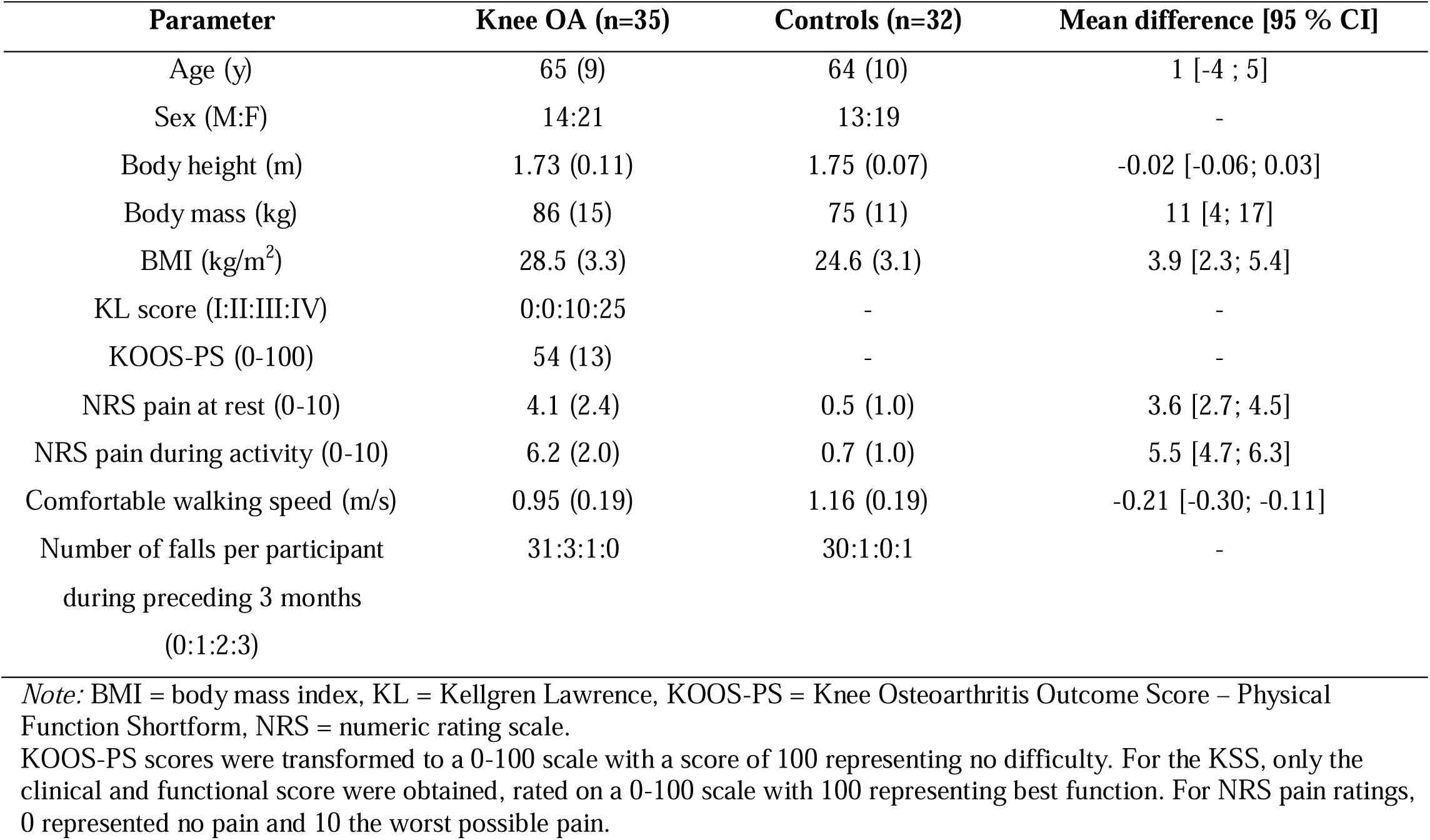
Baseline characteristics of both study groups.

| Parameter | Knee OA (n=35) | Controls (n=32) | Mean difference [95 % CI] |
| --- | --- | --- | --- |
| Age (y) | 65 (9) | 64 (10) | 1 [-4 ; 5] |
| Sex (M:F) | 14:21 | 13:19 | - |
| Body height (m) | 1.73 (0.11) | 1.75 (0.07) | -0.02 [-0.06; 0.03] |
| Body mass (kg) | 86 (15) | 75 (11) | 11 [4; 17] |
| BMI (kg/m<sup>2</sup>) | 28.5 (3.3) | 24.6 (3.1) | 3.9 [2.3; 5.4] |
| KL score (I:II:III:IV) | 0:0:10:25 | - | - |
| KOOS-PS (0-100) | 54 (13) | - | - |
| NRS pain at rest (0-10) | 4.1 (2.4) | 0.5 (1.0) | 3.6 [2.7; 4.5] |
| NRS pain during activity (0-10) | 6.2 (2.0) | 0.7 (1.0) | 5.5 [4.7; 6.3] |
| Comfortable walking speed (m/s) | 0.95 (0.19) | 1.16 (0.19) | -0.21 [-0.30; -0.11] |
| Number of falls per participant during preceding 3 months (0:1:2:3) | 31:3:1:0 | 30:1:0:1 | - |

*Note:* BMI = body mass index, KL = Kellgren Lawrence, KOOS-PS = Knee Osteoarthritis Outcome Score – Physical Function Shortform, NRS = numeric rating scale. KOOS-PS scores were transformed to a 0-100 scale with a score of 100 representing no difficulty. For the KSS, only the clinical and functional score were obtained, rated on a 0-100 scale with 100 representing best function. For NRS pain ratings, 0 represented no pain and 10 the worst possible pain.

We had missing data for one individual with knee OA during the ML perturbations, and for 4 individuals with knee OA during AP perturbation trials. Reasons for missing data were: unable to complete the task due to pain or physical impairment (ML: n=1; AP: n=2), fear (n=1, AP), and lack of time (n=1, AP). Although these participants did not report any falls in the preceding 3 months, their KOOS-PS (range: 38-54) and NRS pain scores during rest (range: 7-9) and activity (range: 7-9) were worse than the group average. Furthermore, six trials of individuals with knee OA (inward affected (n=3), slip affected (n=1), trip affected (n=2)) were not analyzed as the handrail was touched during the balance recovery response.

### 3.1 Mediolateral gait perturbations

For inward perturbations, there was no direct effect of the perturbation visible on the XCoM trajectory (Figure 3). Between the first and second step after perturbation, the XCoM moved approximately 0.05 m less laterally, whereas XCoM excursion was markedly higher between the second and third step after perturbation. XCoM trajectories overlapped between individuals with knee OA and healthy participants. No between-group differences in stepping responses to inward perturbations were found (Table 2 & Figure 4). In both groups, step width decreased with 0.09 m at step 1 and step 2 compared to baseline. This resulted in a decrease in ML MoS of 0.03 m (95% CI: 0.02, 0.04) in the first step, and an increase of 0.01 m (95%: 0.01, 0.02) in the second step compared to baseline. Step time and step length changes after inward perturbations were significant, but small (i.e. 0.01 m longer step lengths in the first step after perturbation and 0.01s shorter step times in the second step after perturbation; Table 2).

**Figure 3:**
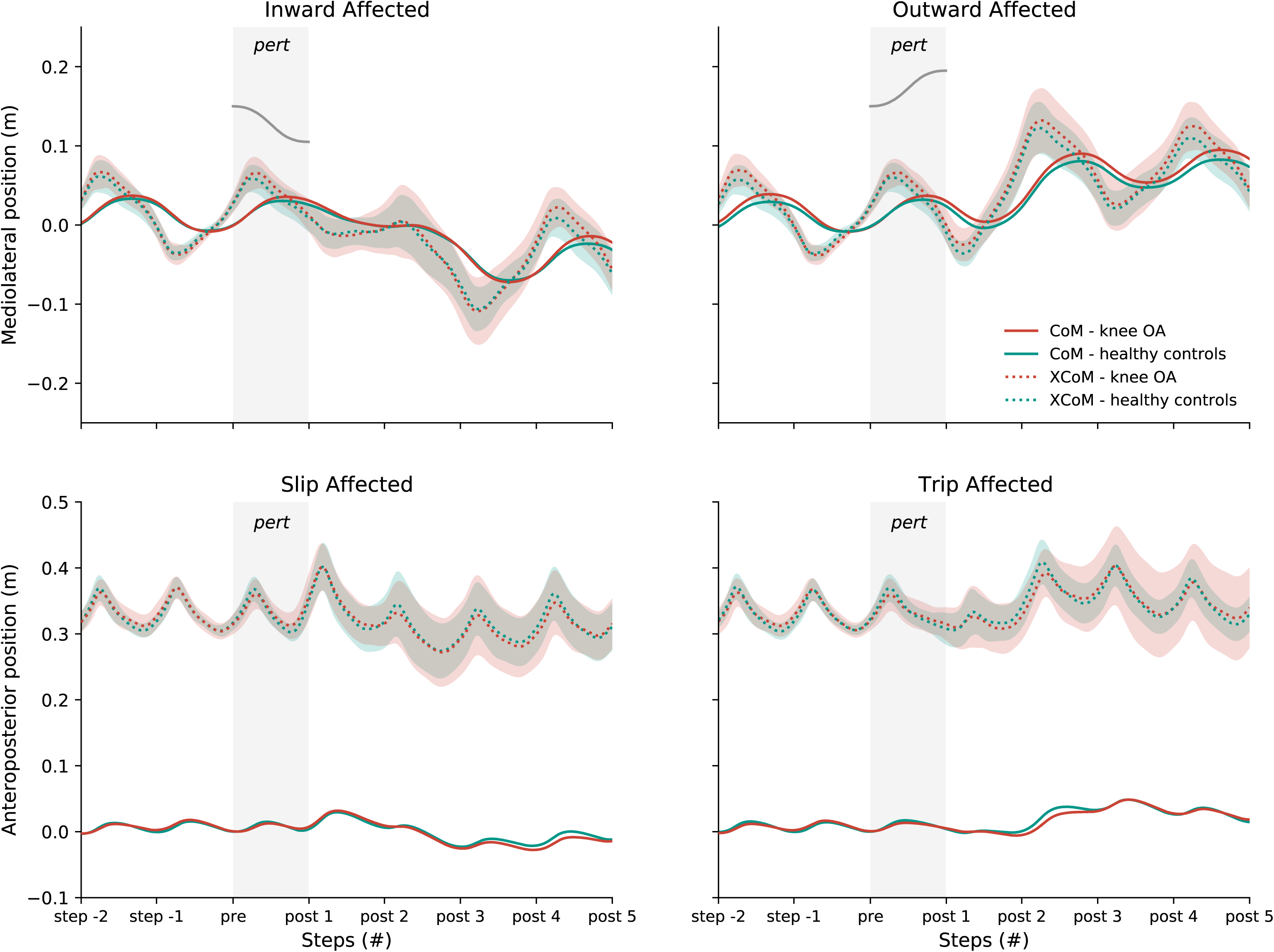
Trajectories of mean center of mass (CoM) and extrapolated center of mass (XCoM) from two steps before until five steps after gait perturbations. Mean values are indicated by the solid and dotted lines. Shaded areas around the extrapolated XCoM represent the standard deviation. Duration of the perturbation (‘pert’) is highlighted by the grey area. For mediolateral perturbations, belt displacement is also indicated by a black line within the grey area.

**Figure 4:**
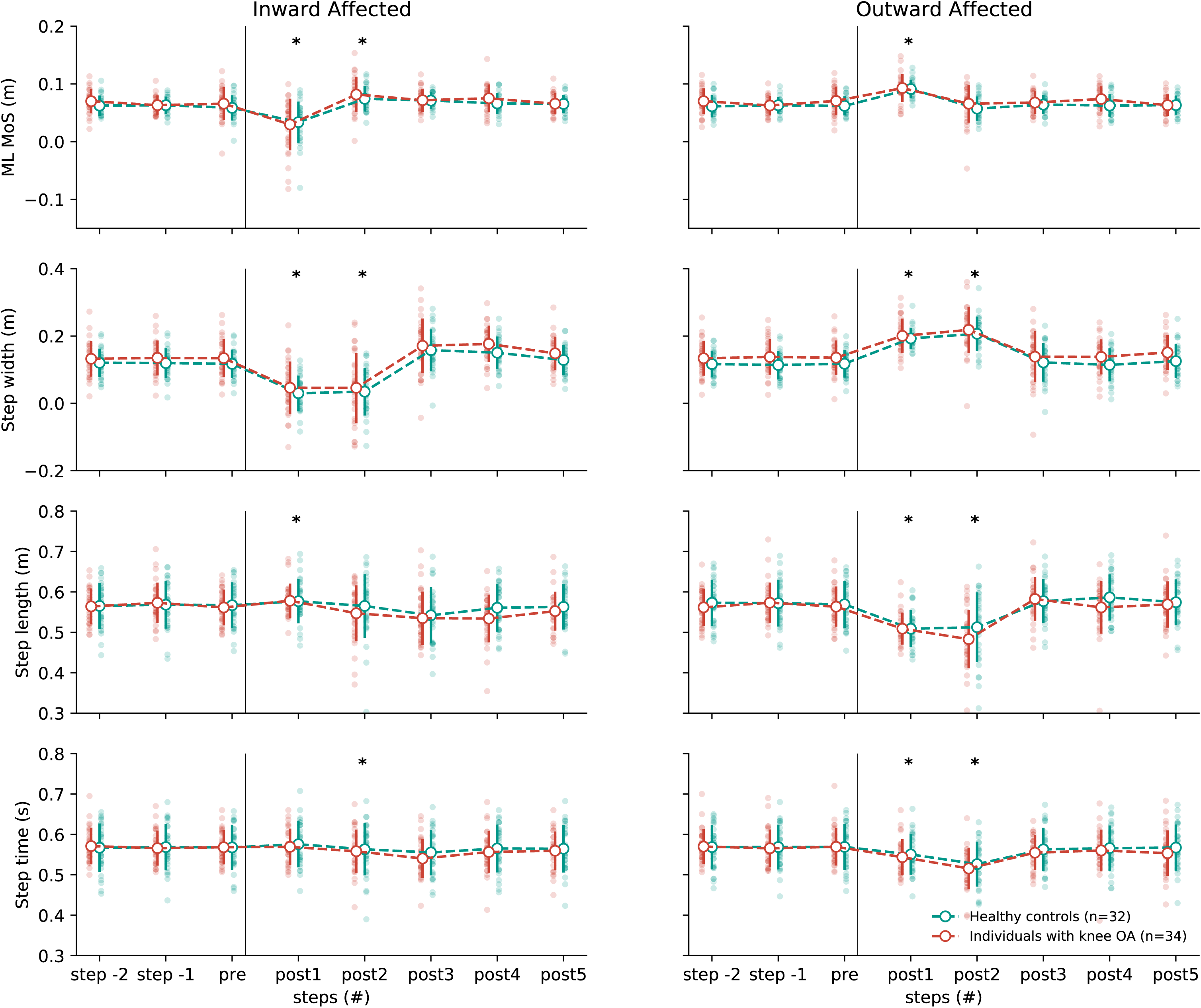
Discrete gait parameters before and after mediolateral gait perturbations. Mean values are indicated by the large white dots, with error bars reflecting the standard deviation. Individual observations are shown with larger transparency. The instance of perturbation is indicated by the black vertical line. Steps before perturbation (i.e. step -2 & step -1) were combined into a baseline score for statistical analysis. *Note:* * significantly different from baseline

**Table 2:** Output of the linear regression models for ML gait perturbations.

| Parameter |  | *Inward affected* |  |  | *Outward affected* |  |  |
| --- | --- | --- | --- | --- | --- | --- | --- |
|  |  | Group Effect |  | Estimated delta score | Group Effect |  | Estimated delta score |
| | | $\beta$ (95% CI) | P-value | | $\beta$ (95% CI) | P-value | |
| ML MoS (m) | Post 1 | -0.01 (-0.03, 0.01) | 0.282 | -0.03 (-0.04, -0.02) | -0.00 (-0.01, 0.01) | 0.790 | 0.03 (0.02, 0.03) |
|  | Post 2 | 0.00 (-0.01, 0.01) | 0.579 | 0.01 (0.01, 0.02) | 0.00 (-0.01, 0.01) | 0.674 | -0.00 (-0.01, 0.00) |
| Step width (m) | Post 1 | -0.00 (-0.02, 0.01) | 0.866 | -0.09 (-0.10, -0.08) | -0.01 (-0.02, 0.01) | 0.263 | 0.07 (0.06, 0.08) |
|  | Post 2 | -0.01 (-0.04, 0.02) | 0.565 | -0.09 (-0.10, -0.07) | -0.01 (-0.03, 0.01) | 0.436 | 0.09 (0.08, 0.10) |
| Step length (m) | Post 1 | -0.00 (-0.01, 0.01) | 0.971 | 0.01 (0.00, 0.01) | 0.00 (-0.01, 0.02) | 0.639 | -0.06 (-0.07, -0.06) |
|  | Post 2 | -0.02 (-0.04, 0.00) | 0.096 | -0.01 (-0.02, 0.00) | 0.02 (-0.05, 0.00) | 0.060 | -0.07 (-0.08, -0.06) |
| Step time (s) | Post 1 | -0.01 (-0.02, 0.00) | 0.091 | 0.00 (-0.00, 0.01) | 0.01 (-0.01, 0.00) | 0.130 | -0.02 (-0.03, -0.02) |
|  | Post 2 | -0.01 (-0.02, 0.01) | 0.404 | -0.01 (-0.01, -0.00) | -0.01 (-0.03, 0.00) | 0.192 | -0.05 (-0.05, -0.04) |

Data are presented as mean difference (95% CI).

Similar to inward perturbations, no instantaneous effect of outward perturbations on the XCoM trajectory was observed. Between the first and second step after perturbation, the XCoM travelled approximately 0.05 m more laterally in both groups (Figure 3). XCoM trajectories were comparable between the two groups. There were no between-group differences in stepping responses to outward perturbations (Table 2). On average, step width increased in the first (mean diff = 0.07 m, 95% CI: 0.06, 0.08) and second step (mean diff = 0.09 m, 95% CI: 0.08, 0.10) after perturbation. Both for individuals with knee OA and healthy participants, ML MoS was 0.03 m (95% CI: 0.02, 0.03) larger than baseline in the first step after outward perturbation, but was not different from baseline in the second step (mean diff = -0.00 m, 95% CI: -0.01, 0.00). Compared to baseline, step length was 0.06 m (95% CI: 0.06, 0.07) shorter in the first step after outward perturbations and 0.07 m (95% CI: 0.06, 0.08) in the second step. Step time was shorter in the first (mean diff = -0.02 s, 95% CI: -0.02, - 0.03) and second (mean diff = -0.05 s, 95% CI: -0.04, -0.05) step after outward perturbations compared to baseline.

### 3.2 Anteroposterior perturbations

Slip perturbations did not have an immediate effect on the XCoM trajectory. Between the first and second step after perturbation, however, the XCoM moved more anteriorly (Figure 3), with both groups showing overlapping XCoM trajectories. In response to the slip perturbation, participants predominantly changed their step length and step time (Figure 5). In the first step after perturbations, step length was 0.10 m (95% CI: 0.09, 0.10) longer compared to baseline in both groups. Step time was 0.02 s (95% CI: 0.02, 0.02) shorter than baseline in both groups. Compared to baseline, AP MoS was 0.05 m (95%: 0.04, 0.05) lower in the first step, followed by a 0.03 m (95% CI: 0.02, 0.04) higher AP MoS in the second step. Changes in AP MoS after slip perturbations were similar between individuals with knee OA and healthy individuals (Table 3). At the second step after perturbation, there was a significant group effect on changes in step length (p=0.045) and step time (p=0.028). Individuals with knee OA showed a 0.02 m (95% CI: 0.00, 0.04) larger decrease in step length compared to baseline, and a 0.02 s (95% CI: 0.00, 0.03) larger reduction in step time. Changes from baseline on step width were small and did not differ between the groups (Table 2).

**Figure 5:**
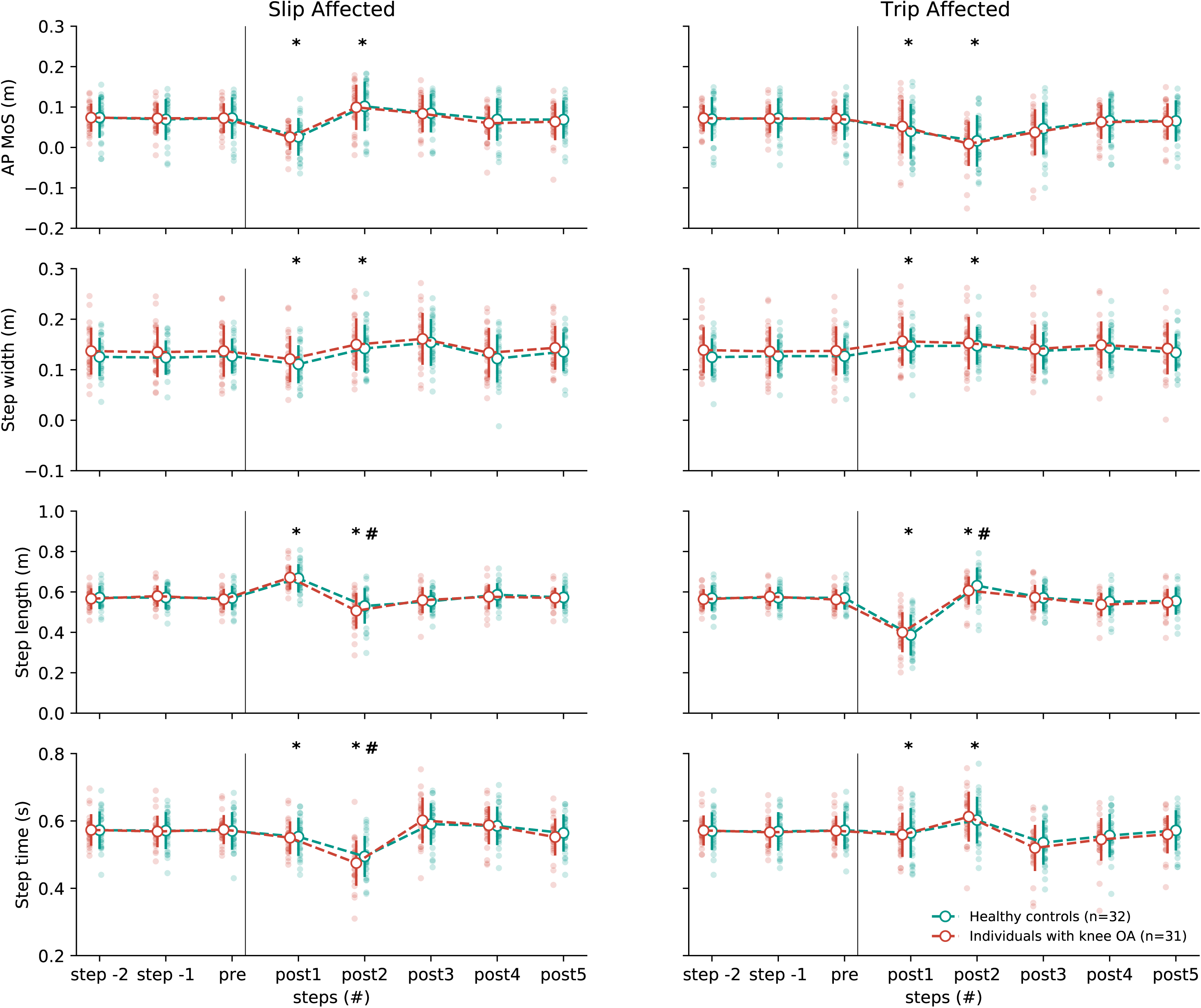
Discrete gait parameters before and after anteroposterior gait perturbations.. Mean values are indicated by the large white dots, with error bars reflecting the standard deviation. Individual observations are shown with larger transparency. The instance of perturbation is indicated by the black vertical line. Steps before perturbation (i.e. step -2 & step -1) were combined into a baseline score for statistical analysis. Note: * significantly different from baseline, # significantly different between groups.

**Table 3:**
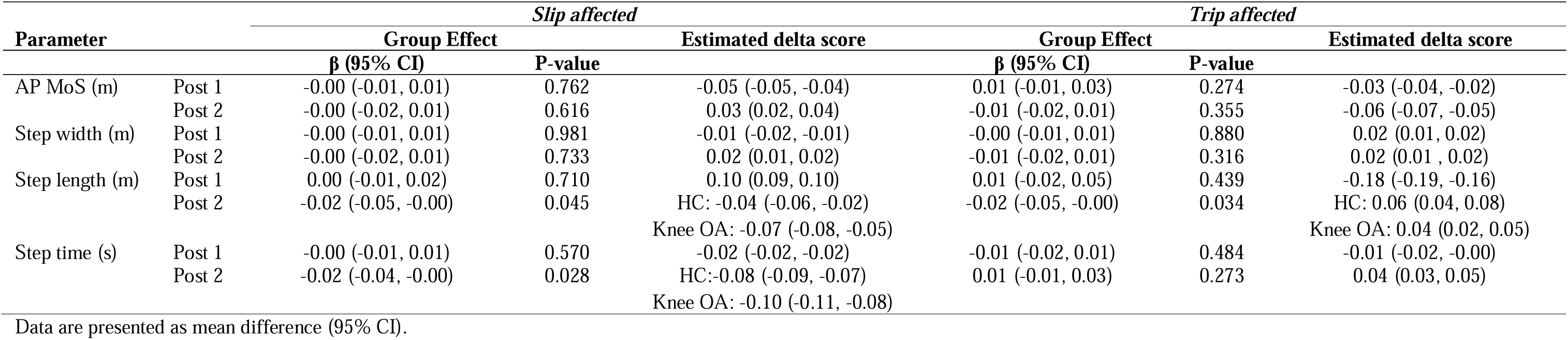
Output of the linear regression models for AP gait perturbations.

Trip perturbations attenuated the forward movement of the XCoM during the first recovery step. Consequently the XCoM was relatively more posterior at the first and second step after perturbation (Figure 3). There were no differences between groups in XCoM trajectory, although the standard deviation of the XCoM trajectory after trip perturbation seemed to be larger in individuals with knee OA. Trip perturbations resulted in a lower step length (mean diff = -0.18 m, 95% CI: -0.19, -0.16) in the first step after perturbation. In the second step after trip perturbation, there was a significant group effect on step length (p=0.034). Step length was 0.06 m (95% CI: 0.04, 0.08) higher in healthy individuals compared to baseline, whereas this was 0.04 m (95% CI: 0.02, 0.05) for individuals with knee OA. Compared to baseline, step time was 0.01 s (95% 0.00, 0.02) shorter in the first step after trip perturbations, and 0.04 s (95% CI: 0.02, 0.05) longer in the second step. There were no group effects on AP MoS in the first and second steps after perturbation (Table 3). For both groups, AP MoS was 0.03 m (95% CI: 0.02, 0.04) lower in the first step after trip perturbations, and 0.06 m (95%: 0.05, 0.07) m lower in the second step. Similar to slip perturbations, the effects of trip perturbations on step width were small and did not differ between the groups (Table 3).

## 4. Discussion

In this study we compared reactive stepping responses to ML and AP gait perturbations between individuals with end-stage knee OA and their healthy peers. After inward as well as outward ML perturbations, individuals with knee OA showed very comparable reactive stepping responses to healthy individuals. In both groups, slip perturbations resulted in a lower AP MoS, and longer step lengths with shorter step times during the first step after perturbation. In the second step after slip perturbation, there was a decrease in step length and step time, which was marginally larger in individuals with knee OA than in healthy individuals. Trip perturbations resulted in a lower AP MoS, and shorter steps with shorter step times in the first step after perturbation in both groups. This initial response was followed by longer steps with longer step times in the second step after perturbation, with individuals with knee OA showing a slightly smaller increase in step length.

Thus, in contrast to our hypothesis, we did not find convincing evidence for impaired reactive stepping responses to gait perturbations in individuals with end-stage knee OA. None of the perturbation modes resulted in group differences in MoS, which was our main outcome of interest. Although it could be argued that taking relatively faster and shorter steps to regain stability – as we found after AP perturbations – may be indicative of poorer balance control (44), these group differences were very small (i.e. 2 cm for step length and 0.02 s for step time). Two main explanations for minor differences between groups can be postulated. To begin with, individuals with knee OA in our study may not have had gait instability, or had only minor localized impairments that they effectively compensated for. Alternatively, our experimental paradigm may not have been challenging enough to trigger large enough balance threats and elucidate instability in the knee OA group. Both options are discussed below.

Given that knee OA leads to a reduced number of mechanoreceptors in the knee capsule and ligaments (45), reduced proprioception (3), lower quadriceps strength (1), and pain, it would be expected that individuals with knee OA have poorer stability than healthy older adults. While postural sway during quiet standing was indeed higher in individuals with knee OA (46, 47), and local dynamic stability tended to be lower during unperturbed walking when compared to healthy adults (5), these reported differences were relatively small. Moreover, it is yet unclear if deviations in these type of balance metrics translate to problems with recovery from external perturbations. So far, studies investigating responses to perturbations in individuals with knee OA have shown mixed results (24–29). For example, Schrijvers *et al.* found larger knee flexion angles and increased co-contraction after AP perturbations in individuals with knee OA with self-reported instability (25). Pater *et al.* found a less optimal recovery strategy from trips over an obstacle during overground walking in individuals with mild to moderate knee OA compared to their healthy peers (27), although the number of fallers after perturbation was similar between groups. In contrast, Kumar *et al.* (28) and Baker *et al.* (29) found no effect of moderate to severe knee OA on change in knee muscle activation and knee kinematics after ML gait perturbations (i.e. 5.8 cm and 3 cm, respectively). Interestingly, none of these studies focused on whole body movement. It may thus well be that individuals with knee OA use adaptations in knee joint kinematics and muscle activation to achieve similar reactive stepping responses as healthy individuals. Moreover, to overcome poorer proprioception due to knee OA, the redundancy of afferent input to and processing within the sensorimotor control system can be exploited (28, 48). By using sensory reweighting, individuals with knee OA may rely more on somatosensory information from other, unaffected structures (49). In light of our results, dynamic balance control may thus still be maintained in individuals with knee OA. Our observation that – in this study – fall rates of individuals with knee OA were relatively low and comparable to healthy individuals further supports that individuals with knee OA in this study may not have had gait stability problems.

A second explanation for the absence of evident instability in the knee OA group could be that the perturbation was insufficiently destabilizing. That is, the ML and AP MoS values before onset of the perturbations in both study groups were higher than (or close to) the perturbed distance (i.e. 4.5 cm for ML perturbations and 12.5 cm for AP perturbations), indicating that there was already some room to cope with these perturbations at baseline. Since the current perturbations were relatively well tolerated by individuals with knee OA, a larger intensity perturbation with potentially better discriminatory capacity may have been feasible. Despite this point, our perturbation paradigm led to clear adaptations in the gait pattern, suggesting that it did challenge the sensorimotor control system. Moreover, responses to these perturbations seem to be robust, as they were comparable to perturbation responses of healthy young (12, 15, 50, 51) and older adults (15, 50) in previous studies with very similar paradigms. Although it might be expected that these paradigms would result in different responses in groups with evident balance problems, this is not yet confirmed in the literature.

This study had a number of limitations that warrant mentioning. First, standardization of walking speed may have led to unnatural walking behavior in some participants as well as differences in experienced difficulty between study groups. Nonetheless, standardization was necessary to separate a potentially confounding influence of walking speed from the effects of knee OA on reactive stepping responses. Moreover, the fixed walking speed was very close to the comfortable walking speed of individuals with knee OA. Secondly, our sample of individuals with unilateral, end-stage knee OA who were scheduled for cruciate retaining total knee arthroplasty may not be representative of all individuals with knee OA. Given that our study group was relatively active, did not have complaints in other joints, and fall rates were low, generalization of our results to the whole knee OA population should be done cautiously.

## 5. Conclusions

Despite considerable knee pain and structural damage to the knee joint, whole-body responses to gait perturbations in individuals with knee OA were not substantially different from healthy individuals. Our results indicate that gait stability in people with knee OA is relatively unimpaired.

## Data Availability

All data produced in the present work are contained in the manuscript

## Acknowledgements

We want to thank Saskia Susan and Jolanda Rubrech for their contribution to patient recruitment and data management, Bart Nienhuis for his contribution to experimental design, and Steven Teerenstra for his advice on statistical analysis.

## References

1. Hoops ML, Rosenblatt NJ, Hurt CP, Crenshaw J, Grabiner MD. Does Lower Extremity Osteoarthritis Exacerbate Risk Factors for Falls in Older Adults? Women’s Health. 2012;8(6):685–98.

2. Shakoor N, Agrawal A, Block JA. Reduced lower extremity vibratory perception in osteoarthritis of the knee. Arthritis Rheum. 2008;59(1):117–21.

3. Knoop J, Steultjens MP, van der Leeden M, van der Esch M, Thorstensson CA, Roorda LD, et al. Proprioception in knee osteoarthritis: a narrative review. Osteoarthritis Cartilage. 2011;19(4):381–8.

4. Hortobágyi T, Garry J, Holbert D, Devita P. Aberrations in the control of quadriceps muscle force in patients with knee osteoarthritis. Arthritis Rheum. 2004;51(4):562–9.

5. Mahmoudian A, Bruijn SM, Yakhdani HRF, Meijer OG, Verschueren SMP, van Dieen JH. Phase-dependent changes in local dynamic stability during walking in elderly with and without knee osteoarthritis. Journal of Biomechanics. 2016;49(1):80–6.

6. Ng CT, Tan MP. Osteoarthritis and falls in the older person. Age and Ageing. 2013;42(5):561–6.

7. Prieto-Alhambra D, Nogues X, Javaid MK, Wyman A, Arden NK, Azagra R, et al. An increased rate of falling leads to a rise in fracture risk in postmenopausal women with self-reported osteoarthritis: a prospective multinational cohort study (GLOW). Annals of the Rheumatic Diseases. 2013;72(6):911.

8. Smith TO, Higson E, Pearson M, Mansfield M. Is there an increased risk of falls and fractures in people with early diagnosed hip and knee osteoarthritis? Data from the Osteoarthritis Initiative. Int J Rheum Dis. 2018;21(6):1193–201.

9. van Schoor NM, Dennison E, Castell MV, Cooper C, Edwards MH, Maggi S, et al. Clinical osteoarthritis of the hip and knee and fall risk: The role of low physical functioning and pain medication. Semin Arthritis Rheum. 2020;50(3):380–6.

10. Doré AL, Golightly YM, Mercer VS, Shi XA, Renner JB, Jordan JM, et al. Lower-extremity osteoarthritis and the risk of falls in a community-based longitudinal study of adults with and without osteoarthritis. Arthritis Care Res (Hoboken). 2015;67(5):633–9.

11. Bruijn SM, Meijer OG, Beek PJ, van Dieën JH. Assessing the stability of human locomotion: a review of current measures. Journal of The Royal Society Interface. 2013;10(83):20120999.

12. Afschrift M, Pitto L, Aerts W, van Deursen R, Jonkers I, De Groote F. Modulation of gluteus medius activity reflects the potential of the muscle to meet the mechanical demands during perturbed walking. Scientific Reports. 2018;8(1):11675.

13. McCrum C, Karamanidis K, Grevendonk L, Zijlstra W, Meijer K. Older adults demonstrate interlimb transfer of reactive gait adaptations to repeated unpredictable gait perturbations. Geroscience. 2020;42(1):39–49.

14. Vlutters M, van Asseldonk EH, van der Kooij H. Center of mass velocity-based predictions in balance recovery following pelvis perturbations during human walking. J Exp Biol. 2016;219(Pt 10):1514–23.

15. Roeles S, Rowe PJ, Bruijn SM, Childs CR, Tarfali GD, Steenbrink F, et al. Gait stability in response to platform, belt, and sensory perturbations in young and older adults. Med Biol Eng Comput. 2018;56(12):2325–35.

16. van Mierlo M, Ambrosius JI, Vlutters M, van Asseldonk EHF, van der Kooij H. Recovery from sagittal-plane whole body angular momentum perturbations during walking. Journal of Biomechanics. 2022;141:111169.

17. van den Bogaart M, Bruijn SM, van Dieën JH, Meyns P. The effect of anteroposterior perturbations on the control of the center of mass during treadmill walking. Journal of Biomechanics. 2020;103:109660.

18. Hof AL, Gazendam MGJ, Sinke WE. The condition for dynamic stability. Journal of Biomechanics. 2005;38(1):1–8.

19. O’Connor SM, Kuo AD. Direction-dependent control of balance during walking and standing. J Neurophysiol. 2009;102(3):1411–9.

20. Collins SH, Kuo AD. Two independent contributions to step variability during over-ground human walking. PLoS One. 2013;8(8):e73597.

21. McGeer T. Passive Dynamic Walking. The International Journal of Robotics Research. 1990;9(2):62–82.

22. van Leeuwen M, Bruijn S, van Dieën J. Mechanisms that stabilize human walking. Brazilian Journal of Motor Behavior. 2022;16(5):326–51.

23. Bruijn SM, van Dieën JH. Control of human gait stability through foot placement. J R Soc Interface. 2018;15(143).

24. Ren X, Lutter C, Kebbach M, Bruhn S, Yang Q, Bader R, et al. Compensatory Responses During Slip-Induced Perturbation in Patients With Knee Osteoarthritis Compared With Healthy Older Adults: An Increased Risk of Falls? Frontiers in Bioengineering and Biotechnology. 2022;10.

25. Schrijvers JC, van den Noort JC, van der Esch M, Harlaar J. Neuromechanical assessment of knee joint instability during perturbed gait in patients with knee osteoarthritis. Journal of Biomechanics. 2021;118:110325.

26. Elkarif V, Kandel L, Rand D, Schwartz I, Greenberg A, Portnoy S. Kinematics following gait perturbation in adults with knee osteoarthritis: Scheduled versus not scheduled for knee arthroplasty. Gait & Posture. 2020;81:144–52.

27. Pater ML, Rosenblatt NJ, Grabiner MD. Knee osteoarthritis negatively affects the recovery step following large forward-directed postural perturbations. Journal of Biomechanics. 2016;49(7):1128–33.

28. Kumar D, Swanik C, Reisman DS, Rudolph KS. Individuals with medial knee osteoarthritis show neuromuscular adaptation when perturbed during walking despite functional and structural impairments. Journal of Applied Physiology. 2013;116(1):13–23.

29. Baker M, Stanish W, Rutherford D. Walking challenges in moderate knee osteoarthritis: A biomechanical and neuromuscular response to medial walkway surface translations. Human Movement Science. 2019;68:102542.

30. McCrum C, Willems P, Karamanidis K, Meijer K. Stability-normalised walking speed: A new approach for human gait perturbation research. Journal of Biomechanics. 2019;87:48–53.

31. Orendurff MS, Segal AD, Klute GK, Berge JS, Rohr ES, Kadel NJ. The effect of walking speed on center of mass displacement. J Rehabil Res Dev. 2004;41(6a):829–34.

32. Boekesteijn RJ, van Gerven J, Geurts ACH, Smulders K. Objective gait assessment in individuals with knee osteoarthritis using inertial sensors: A systematic review and meta-analysis. Gait & Posture. 2022;98:109–20.

33. Boekesteijn RJ, Keijsers NLW, Defoort K, Mancini M, Bruning FJ, El-Gohary M, et al. Real-world gait and turning in individuals scheduled for total knee arthroplasty. medRxiv. 2023:2023.09.13.23295243.

34. Kellgren JH, Lawrence JS. Radiological assessment of osteo-arthrosis. Ann Rheum Dis. 1957;16(4):494–502.

35. Perruccio AV, Stefan Lohmander L, Canizares M, Tennant A, Hawker GA, Conaghan PG, et al. The development of a short measure of physical function for knee OA KOOS-Physical Function Shortform (KOOS-PS) - an OARSI/OMERACT initiative. Osteoarthritis Cartilage. 2008;16(5):542–50.

36. Insall JN, Dorr LD, Scott RD, Scott WN. Rationale of the Knee Society clinical rating system. Clin Orthop Relat Res. 1989(248):13–4.

37. Lamb SE, Jørstad-Stein EC, Hauer K, Becker C, on behalf of the Prevention of Falls Network E, Outcomes Consensus G. Development of a Common Outcome Data Set for Fall Injury Prevention Trials: The Prevention of Falls Network Europe Consensus. Journal of the American Geriatrics Society. 2005;53(9):1618–22.

38. Davis RB, Õunpuu S, Tyburski D, Gage JR. A gait analysis data collection and reduction technique. Human Movement Science. 1991;10(5):575–87.

39. Tisserand R, Robert T, Dumas R, Chèze L. A simplified marker set to define the center of mass for stability analysis in dynamic situations. Gait & Posture. 2016;48:64–7.

40. Hak L, Houdijk H, Beek PJ, van Dieën JH. Steps to Take to Enhance Gait Stability: The Effect of Stride Frequency, Stride Length, and Walking Speed on Local Dynamic Stability and Margins of Stability. PLOS ONE. 2013;8(12):e82842.

41. Boekesteijn RJ, Smolders JMH, Busch VJJF, Geurts ACH, Smulders K. Independent and sensitive gait parameters for objective evaluation in knee and hip osteoarthritis using wearable sensors. BMC Musculoskeletal Disorders. 2021;22(1):242.

42. Zeni JA, Jr., Richards JG, Higginson JS. Two simple methods for determining gait events during treadmill and overground walking using kinematic data. Gait Posture. 2008;27(4):710–4.

43. McLlroy WE, Maki BE. Adaptive changes to compensatory stepping responses. Gait & Posture. 1995;3(1):43–50.

44. Maki BE, McIlroy WE. Control of rapid limb movements for balance recovery: age-related changes and implications for fall prevention. Age and Ageing. 2006;35(suppl_2):ii12–ii8.

45. Çabuk H, Kuşku Çabuk F, Tekin AÇ, Dedeoğlu SS, Çakar M, Büyükkurt CD. Lower numbers of mechanoreceptors in the posterior cruciate ligament and anterior capsule of the osteoarthritic knees. Knee Surgery, Sports Traumatology, Arthroscopy. 2017;25(10):3146–54.

46. Hassan BS, Mockett S, Doherty M. Static postural sway, proprioception, and maximal voluntary quadriceps contraction in patients with knee osteoarthritis and normal control subjects. Annals of the Rheumatic Diseases. 2001;60(6):612.

47. Hinman RS, Bennell KL, Metcalf BR, Crossley KM. Balance impairments in individuals with symptomatic knee osteoarthritis: a comparison with matched controls using clinical tests. Rheumatology. 2002;41(12):1388–94.

48. Barry BK, Sturnieks DL. How important are perturbation responses and joint proprioception to knee osteoarthritis? Journal of Applied Physiology. 2014;116(1):1–2.

49. Mahmoudian A, van Dieen JH, Baert IAC, Jonkers I, Bruijn SM, Luyten FP, et al. Changes in proprioceptive weighting during quiet standing in women with early and established knee osteoarthritis compared to healthy controls. Gait & Posture. 2016;44:184–8.

50. McIntosh EI, Zettel JL, Vallis LA. Stepping Responses in Young and Older Adults Following a Perturbation to the Support Surface During Gait. J Mot Behav. 2017;49(3):288–98.

51. Li J, Huang HJ. Small directional treadmill perturbations induce differential gait stability adaptation. Journal of Neurophysiology. 2021;127(1):38–55.

